# Immunophenotyping and machine learning identify distinct immunotypes that predict COVID-19 clinical severity

**DOI:** 10.1101/2021.05.07.21256531

**Authors:** Yvonne M. Mueller, Thijs J. Schrama, Rik Ruijten, Marco W.J. Schreurs, Dwin G.B. Grashof, Harmen J. G. van de Werken, Daniel Alvarez de la Sierra, Caoimhe H. Kiernan, Melisa D. Castro Eiro, Marjan van Meurs, Inge Brouwers-Haspels, Manzhi Zhao, Ling Li, Harm de Wit, Christos A. Ouzounis, Merel E. P. Wilmsen, Tessa Alofs, Danique A. Laport, Tamara van Wees, Geoffrey Kraker, Maria C. Jaimes, Sebastiaan Van Bockstael, Manuel Hernández-González, Casper Rokx, Bart J.A. Rijnders, Ricardo Pujol-Borrell, Peter D. Katsikis

**Author notes:** Address correspondence to Peter D. Katsikis.

## Abstract

Quantitative or qualitative differences in immunity may drive and predict clinical severity in COVID-19. We therefore measured modules of serum pro-inflammatory, anti-inflammatory and anti-viral cytokines in combination with the anti-SARS-CoV-2 antibody response in COVID-19 patients admitted to tertiary care. Using machine learning and employing unsupervised hierarchical clustering, agnostic to severity, we identified three distinct immunotypes that were shown post-clustering to predict very different clinical courses such as clinical improvement or clinical deterioration. Immunotypes did not associate chronologically with disease duration but rather reflect variations in the nature and kinetics of individual patient’s immune response. Here we demonstrate that immunophenotyping can stratify patients to high and low risk clinical subtypes, with distinct cytokine and antibody profiles, that can predict severity progression and guide personalized therapy.

## Introduction

The newly emerged SARS-CoV-2 virus has caused COVID-19 pandemic and infected more than 120 million people over the world, resulting in more than 2.8 million deaths (*1*). In the absence of a highly effective therapy against COVID-19, there remains an urgent need to understand both the pathological mechanisms that lead to severe disease but to also identify clear phenotypes that predict disease severity progression and outcome as this may instruct a more personalized therapy. In an attempt to understand the features of COVID-19 that associate with disease severity, studies have aimed at capturing the perturbation of the immune system and the associated inflammatory syndrome observed. Some of these studies have applied high dimensional analysis using multiplex cytokines, flow or mass cytometry, or scRNAseq to identify changes in cytokine profiles, peripheral blood immune cell composition and/or gene expression related to COVID-19 severity. Universally, however, these studies have employed disease severity classification to identify immunotypes that characterize mild, moderate or severe disease (*2-8*). Although, these studies have identified specific changes present in COVID-19 patients compared with healthy individuals, identifying clear immunotypes that strongly associate with or predict disease severity has proven more challenging (*2-5*). Defining, however, immunotypes based on clinical severity is based on the assumption that a single mechanism underlies all patients and that kinetics are exclusively driven by days of infection. This approach is, thus, hampered by the dynamic nature of the immune and inflammatory response to SARS-CoV-2 virus, the very different kinetics that individual patients may exhibit, and the likelihood that very different immune mechanisms underlie the same clinical severity.

### Distinct immunotypes are identified by machine learning in acute COVID-19 disease

In this study, we chose to take an unbiased approach in terms of clinical severity to identify immunotypes by first defining immunotypes in COVID-19 patients and then examining if these relate to clinical severity and progression. At time of study entry, we measured in the serum of COVID-19 patients (Rotterdam discovery cohort; n=50, Supplementary Table 1) modules of specific cytokines with pro-inflammatory, anti-inflammatory or anti-viral activities. We combined these serum cytokines with the host adaptive antibody response and applied machine learning and unsupervised hierarchical clustering to identify immunophenotypes that capture both innate and adaptive responses to SARS-CoV-2 infection. Importantly, we did not use clinical severity as a clustering parameter. Using this approach, we identified three distinct immunotypes, (labelled: balanced response immunotype: BRI, excessive inflammation immunotype: EXI and low antibody immunotype: LAI) in acutely infected COVID-19 patients (Figure 1A). To validate these immunotypes, we applied the same machine learning approach on a second independent cohort of patients (Barcelona validation cohort; n=88, Supplementary Table 1) (Figure 1B). Principal component analysis (PCA) showed that measurements of the Barcelona cohort data matched very well with the Rotterdam data (Figure 1C). Independent unsupervised hierarchical clustering of the measurements from the Barcelona cohort consistently revealed a very similar classification of patients into 3 distinct immunotypes BRI, EXI and LAI who exhibited similar cytokine and antibody characteristics as those discovered in the Rotterdam cohort (Fig 1A and B).

**Figure 1:**
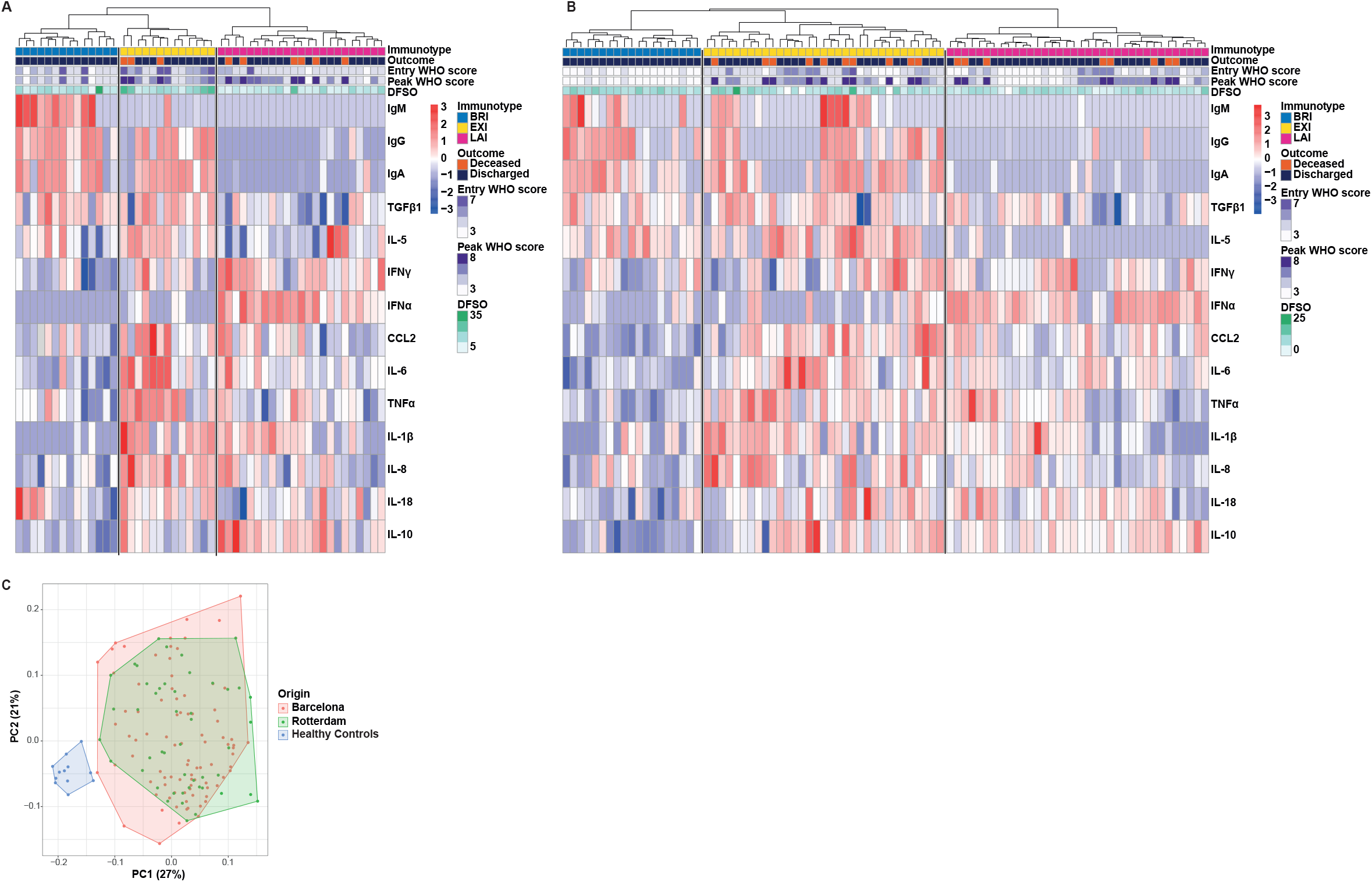
Unsupervised hierarchical clustering identifies three distinct immunotypes in acute COVID-19 patients. Applying machine learning unsupervised hierarchical clustering solely to serum cytokines and anti-SARS-CoV-2 antibodies identifies 3 distinct immunotypes. Analysis was performed on samples collected at study entry and without clinical data input. The three immunotypes, identified independently in two patient cohorts, are depicted in **A)** the heat map of the Rotterdam discovery cohort (n=50) and **B)** the heat map of the Barcelona validation cohort (n=88) using row-based log transformed *z*-scores. **C)** Principal Component Analysis (PCA) of serum cytokines and anti-SARS-CoV-2 antibodies shows that both Rotterdam and Barcelona cohorts cluster together while they lie apart from healthy controls. The first two components with their percentage of variance are shown in parentheses. Red-blue color depicts z-scores. Top banners of heat maps show: immunotypes BRI, EXI and LAI, WHO clinical score at entry and the peak during hospitalization, patient death and discharge, and days from symptoms onset (DFSO).

The 3 immunotypes were characterized by distinct serum cytokine profiles and anti-SARS-CoV-2 antibody responses (Figure 2). Compared to healthy controls, all 3 immunotypes had increased pro-inflammatory cytokines (Figure 2A and Supplementary Figure S1A), displaying further significant differences between them. Immunotype BRI was characterized by low pro-inflammatory, antiviral and anti-inflammatory cytokines and normal TGFβ1 levels (Figure 2A, Supplementary Figure S1A). BRI exhibited robust IgM, IgG and IgA anti-SARS-CoV-2 nucleocapsid protein (N-protein) antibodies (Figure 2B). In contrast, EXI had a much more pro-inflammatory profile, low IFNα and normal TGFβ1 (Figure 2A). Immunotype EXI was also associated with IgM, IgG and IgA anti-SARS-CoV-2 antibodies (Figure 2B). Immunotype LAI exhibited a distinct profile from the previous two and was characterized by the presence of a strong IFNα response, reduced TGFβ1 (Figure 2A), and very low antibody immunity (Figure 2B). Similar immunotype-specific antibody differences were also seen with anti-RDB antibodies and neutralization titers (data not shown). Pro-inflammatory cytokines were significantly higher in EXI compared to both other immunotypes (Fig 2A). IL-17A and IL-5 were increased in only some EXI or LAI patients, very few patients had IL-2 or IL-12 in serum (Supplementary Figure S1A) while IL-4 was undetectable (data not shown). Thus, the signatures of EXI and LAI differed in terms of their anti-SARS-CoV-2 antibody response, the level of pro-inflammatory cytokines and IFNα while TGFβ1 was uniquely downregulated in LAI (Figure 1B and 2).

**Figure 2:**
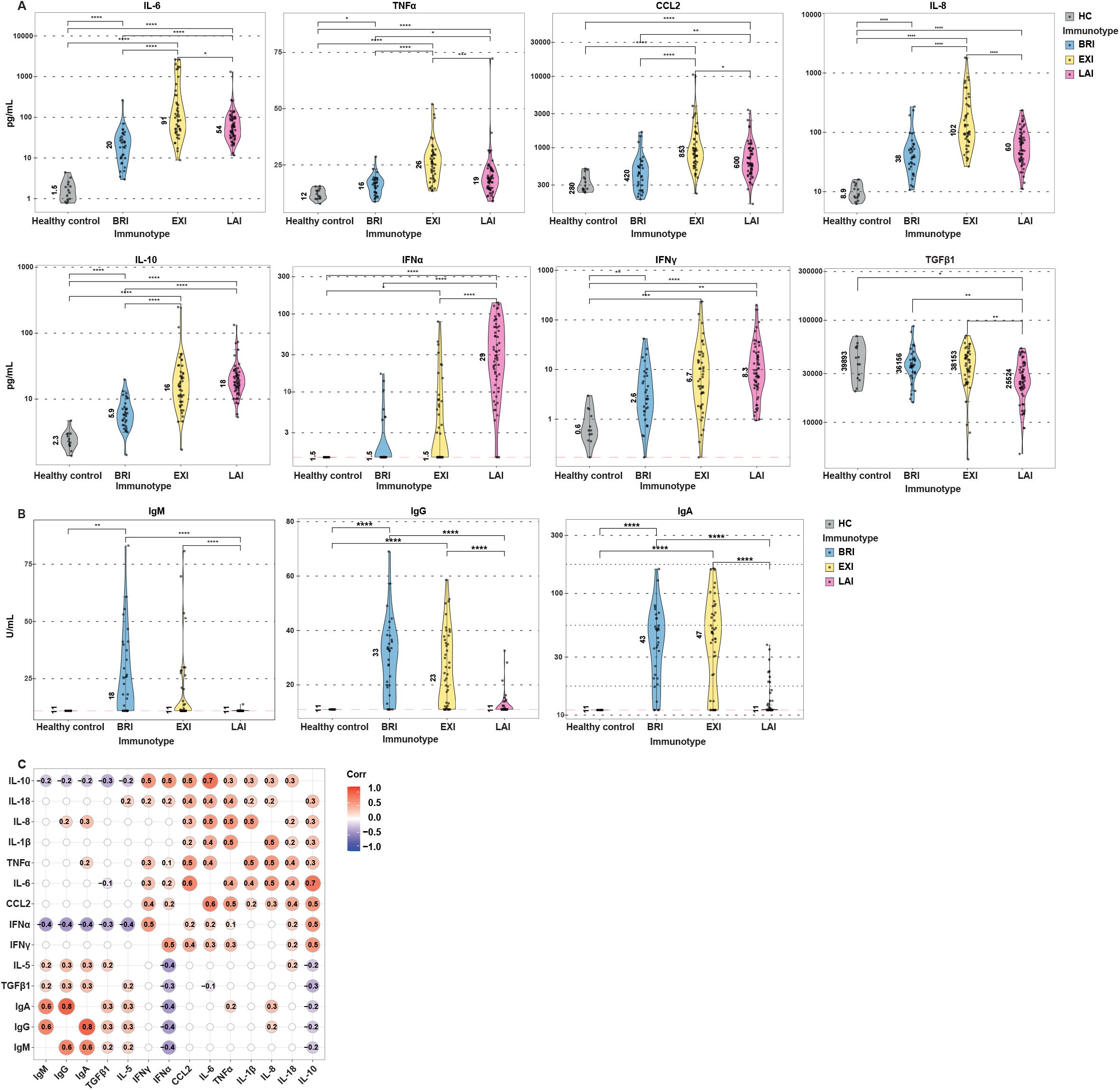
COVID-19 immunotypes have distinct cytokine and anti-SARS-CoV-2 antibody levels. Immunotypes BRI, EXI and LAI have distinct patterns of cytokines and anti-SARS-CoV-2 antibodies. **A)** serum cytokines and **B)** IgM, IgG and IgA anti-SARS-CoV-2 antibodies are shown for BRI, EXI and LAI patients and healthy controls. Violin plots with single data points depict the range of levels and number indicates medians. Wilcoxon rank-sum statistical tests with Bonferroni correction was applied on all six comparisons but only shown for significant differences. Red dashed line indicates assay Limit of Detection. **C)** Correlograms depicting correlations between cytokines and antibodies. Red indicates positive and blue negative correlation. Only statistically significant association (p < 0.05) are shown. Spearman’s rho (R_S_) is shown in circles. *p < 0.05, **p < 0.01, ***p < 0.001, and ****p < 0.0001.

As expected, we found that anti-SARS-CoV-2 IgM, IgG and IgA antibodies correlated strongly with each other (Figure 2C). The strongest cytokine correlation with antibodies, was a negative correlation with IFNα (Figure 2C). This could indicate that antibodies reduce viral loads and thus IFNα or conversely, high IFNα levels delay or inhibit antibody production. Pro-inflammatory cytokines correlated with each other (Figure 2C). Antiviral cytokines IFNα and IFNγ correlated with each other, while IFNα negatively correlated with TGFβ1 (Figure 2C). Thus characteristics of the 3 immunotypes could be driven by distinct cytokine networks in action.

### COVID-19 immunotypes predict clinical severity progression

We next investigated whether these distinct immunotypes associated with clinical parameters (Supplementary Table 2). As mentioned, clinical severity was not used as a parameter for the unsupervised hierarchical clustering. At study entry, BRI and LAI did not differ in WHO clinical severity scores (*9*) while EXI was significantly higher (Figure 3A). Thus at study entry, the immunotypes are not determined by disease severity. To assess clinical severity progression during hospitalization, we examined the highest/worst clinical score that patients exhibited within 30 days of admittance. During hospitalization, EXI and LAI were characterized by clinical deterioration and higher peak WHO clinical severity scores (media peak score of 6 for both) (Figure 3A). In contrast, BRI improved after entry and clinical scores declined (the median peak score of 3, was the score of entry) (Figure 3A). Reflecting the more severe disease scores during hospitalization, all mortality occurred in EXI and LAI patients, while no patients died in BRI (Figure 1A&B and Figure 3B). These differences in severity between immunotypes could not be attributed to age as these did not vary significantly (Figure 3C). Thus despite immunotypes having a relatively mild disease at study entry, the EXI and LAI phenotypes captured patients that would clinically deteriorate after hospitalization, while BRI identified patients that would improve clinically.

**Figure 3:**
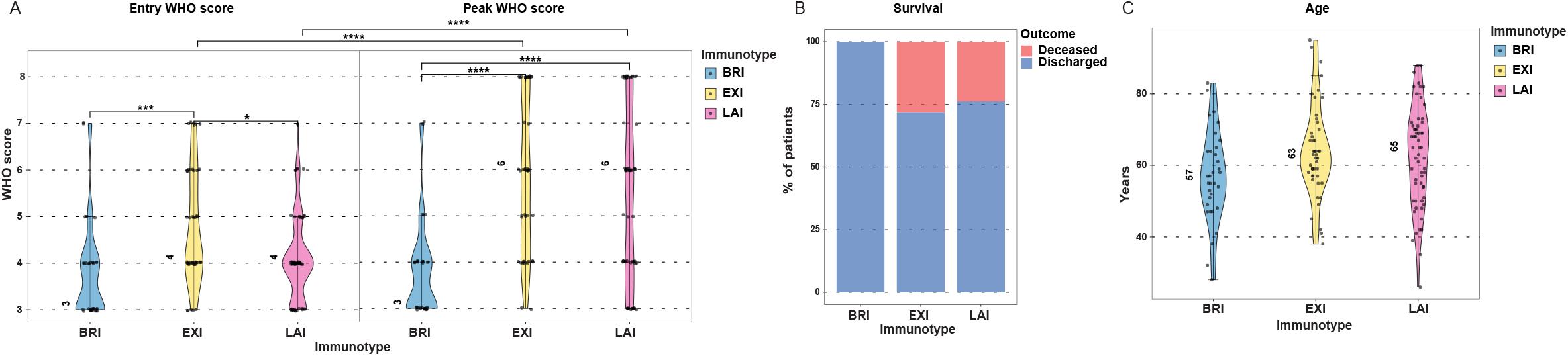
Immunotypes predict clinical improvement or deterioration in acute COVID-19. COVID-19 immunotype BRI identifies at study entry patients that will clinically improve while immunotypes EXI and LAI predict clinical deterioration as peak severity scores during hospitalization are higher than entry scores. Violin plots show **A)** WHO clinical score at study entry and peak WHO clinical score during hospitalization, **B)** Percentage of deceased patients per immunotype and **C)** Age of patients per immunotype. Medians indicated by number in plots. Significance of WHO scores was tested using a Wilcoxon rank-sum tests for entry WHO score and peak WHO score, significant differences in age were tested using Student’s *t*-tests with Bonferroni correction. *p < 0.05, **p < 0.01, ***p < 0.001, and ****p < 0.0001.

### COVID-19 immunotypes are not defined by disease duration

Although the 3 immunotypes differ at time of entry in the number of days from symptoms onset (DFSO), with disease duration shortest in LAI, differences in DFSO did not explain these phenotypes (Figure 4A, B & C). Cytokine and antibody levels did not correlate with DFSO, suggesting that immunotypes are not determined by disease duration (Figure 4B and C). More importantly, the kinetics of antibodies and cytokines differed between the three immunotypes with LAI even after DFSO of 10 still having high levels of IFNα while the antibody responses remained muted (Figure 4B and C). IL-6 and TNFα, on the other hand, were high in EXI patients already at DFSO of 5 days (Figure 4B). For the individual immunotypes, anti-SARS-CoV-2 antibody responses had different trajectories in terms of DFSO. In BRI and EXI patients, antibodies come up early and stay up (Figure 4C). The frequency of patients positive for anti-SARS-CoV-2 antibodies was increased in BRI and EXI compared to LAI irrespective of time since symptoms onset (Figure 4D). Thus time of infection could not explain LAI’s high IFNα or low antibodies. This supports the notion that immunotypes reflect the individual patient’s nature of the response and the rate of development of immunity rather than a strictly linear chronological relationship to duration of infection.

**Figure 4:**
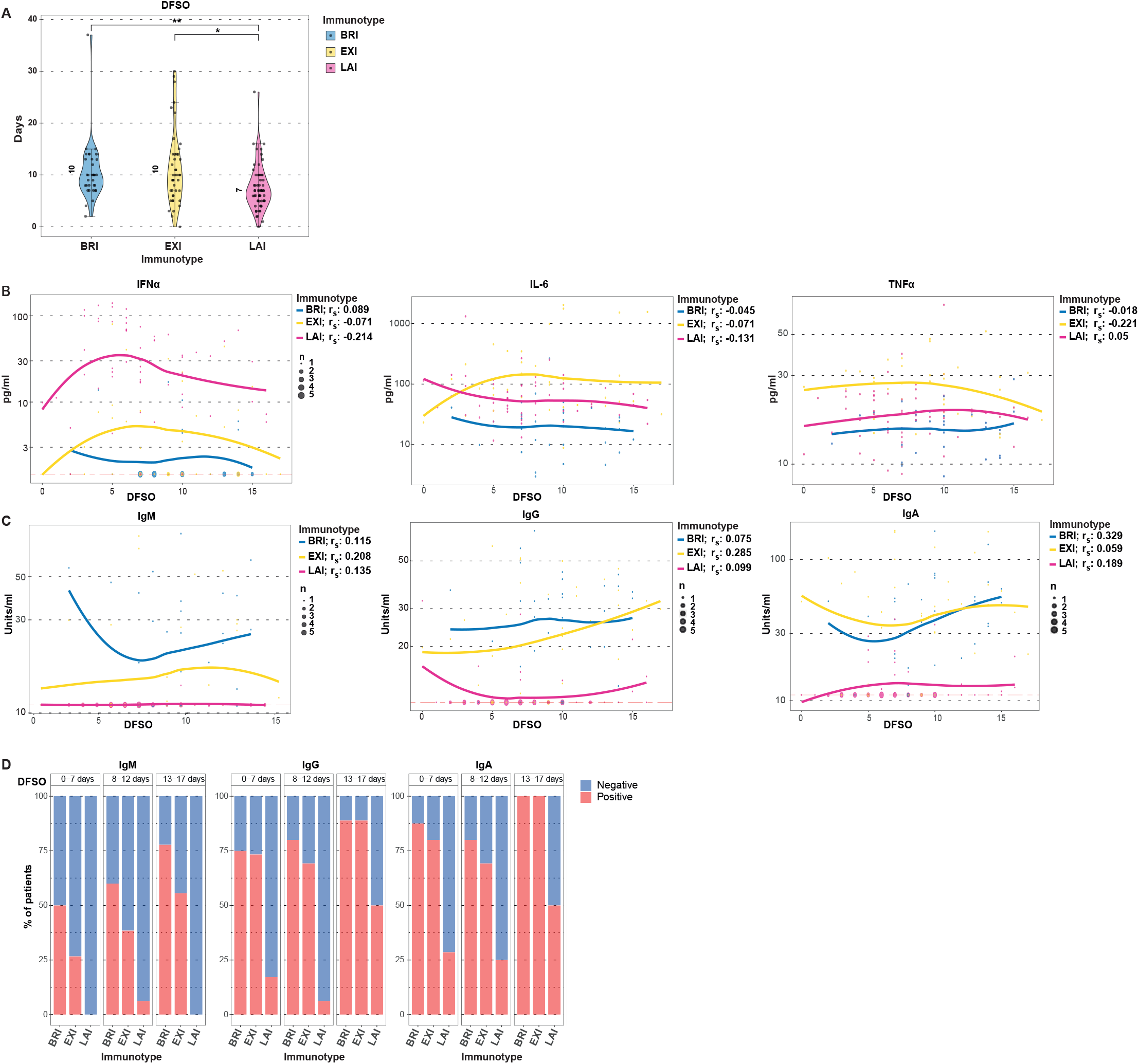
Disease duration does not determine immunotypes. Cytokines differences and the absence of antibodies in LAI are not due to lesser disease duration. **A)** Days from symptoms onset (DFSO). Medians shown by number. Wilcoxon rank-sum tests with Bonferroni correction were applied. *p < 0.05, **p < 0.01. **B)** Correlations of IFNα, IL-6 and TNFα with DFSO per immunotype shown. **C)** Correlations of anti-SARS-CoV-2 antibodies with DFSO is shown per immunotype. For B and C non-parametric local polynomial regression lines plotted using the locally estimated scatterplot smoothing (LOESS) method are shown for individual immunotypes. Spearman’s rho (r_s_) and p value is shown when significant. **D)** The percentage of antibody positivity for the 3 immunotypes is depicted in relation to their DFSO. Red depict percent of patients positive for antibodies, blue depicts percent negative.

### Immunotypes differ in their clinical laboratory characteristics

The 3 immunotypes differed significantly in terms of plasma inflammation markers and blood cell numbers (Supplementary Table 2). Plasma levels of markers of inflammation C-reactive protein (CRP), ferritin, d-dimers, Lactate dehydrogenase (LDH) were all significantly higher in EXI (Figure 5A). Strikingly, these were not increased in LAI compared to BRI. Leukocytes and neutrophils were highest in EXI, while lymphocyte levels trended to be lower in EXI and LAI but were not significant (Figure 5B). Thrombocytes, however, were significantly lower in LAI patients (Figure 5B, Supplementary Table 2). Viral loads in nasopharyngeal swabs were not significantly different between immunotypes but trended to be higher in LAI (Figure 5C), while within LAI patients, viral loads did not correlate with IFNα levels (Figure 5D).

**Figure 5:**
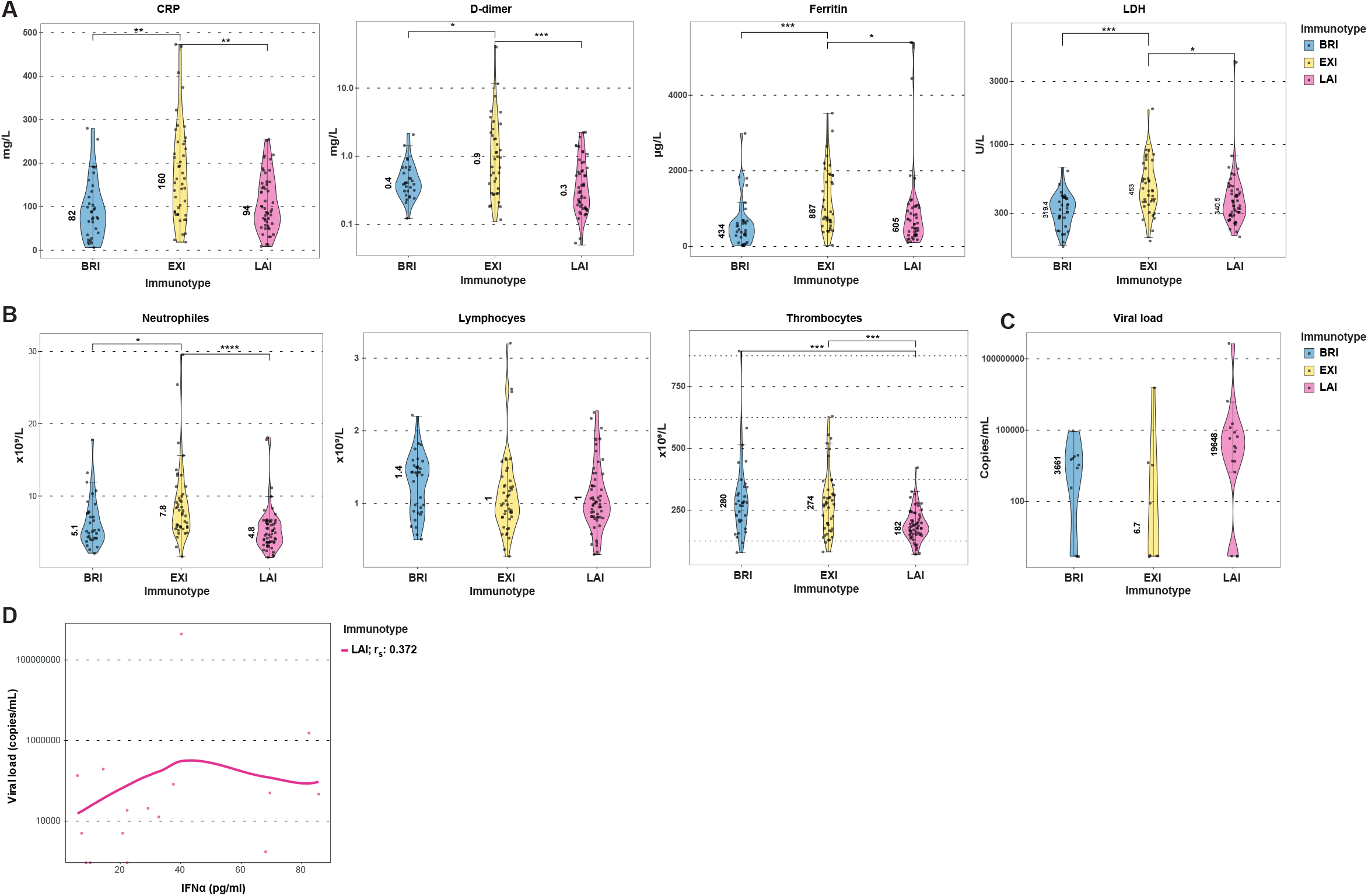
Immunotypes differ in their clinical laboratory characteristics. **A)** CRP, d-dimer, ferritin and LDH measurements shown. **B)** Violin plots of blood neutrophil, lymphocyte and thrombocyte counts. **C)** Viral loads of immunotypes at entry shown. Medians shown by number in plots. **D)** the scatterplot of IFNα and viral loads in LAI patients shown. Line depicts the non-parametric local polynomial regression line plotted using the locally estimated scatterplot smoothing (LOESS) method. Spearman’s rho (r_s_) is indicated. Correlation was not significant. A subgroup of Rotterdam patients with nasopharyngeal viral load measurements are shown. Wilcoxon rank-sum tests with Bonferroni correction were applied for each measurement in A-C. *p < 0.05, **p < 0.01, ***p < 0.001, and ****p < 0.0001.

### Deep phenotyping of immunotypes reveals distinct cellular changes in blood of COVID-19 patients

To acquire insight into the potential mechanisms that are behind the described 3 immunotypes we used 40-color spectral flow cytometry and identified the immune cell subset changes associated with these immunotypes (Figure 6). Although COVID-19 patients as a whole differed clearly from healthy individuals, comparing immunotypes at study entry found remarkably few differences between them. COVID-19 patients compared to healthy individuals had increased plasmablasts (Figure 6B, C and D, Supplementary Table 3). The pro-inflammatory intermediate monocytes 1 and 2 (*10*) and pro-inflammatory IgD negative non-conventional memory B cells (IgD- CD27- B cells) (*11*) were both increased in blood of patients (Figure 6C, Supplementary Figure 2, Supplementary Table 3). In contrast, non-classical monocytes were reduced in blood of patients (Figure 6C, Supplementary Figure 2, Supplementary Table 3), and loss of these anti-inflammatory cells may further contribute to immune activation (*12, 13*). Plasmacytoid DC and conventional DC were also both reduced in patients (Figure 6B, C and D, Supplementary Table 3). When the 3 immunotypes were compared with each other, remarkably few differences existed (Figure 6E and F, Supplementary Table 3). Only intermediate monocytes 2 were strongly increased in EXI compared to LAI (Figure 6F and G). Overall, despite the large differences between COVID-19 patients and healthy controls, the differences between immunotypes were subtle, underscoring a disconnect between peripheral blood cells and systemic plasma cytokines in the immunotypes.

**Figure 6:**
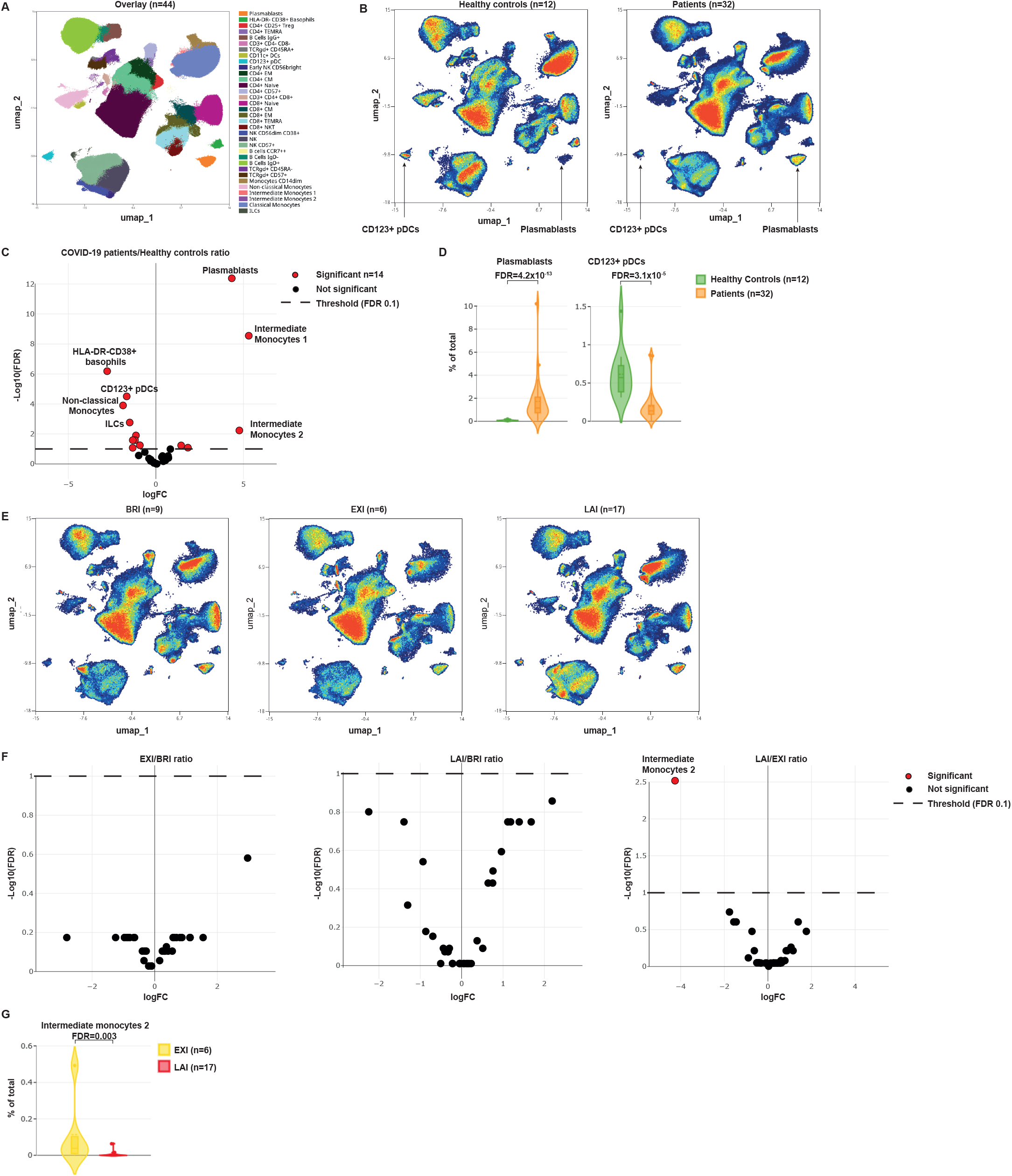
High dimensional flow cytometry reveals distinct immune subset differences between immunotypes. **A)** Population cluster identification in high-dimensional 40-color flow cytometry data using Uniform Manifold Approximation and Projection (UMAP) dimensionality reduction. Visualization is performed on combined COVID-19 patients and healthy control data (n=44). **B)** UMAP plots of healthy controls (n=12) and all COVID-19 patients (n=32). Plasmablast and pDC indicated by arrows. **C)** Volcano plots of the comparison COVID-19 patients/Healthy controls. Positive fold changes indicate cellular population increases in patients while negative fold change indicate decreases in patients. **D)** Plasmablast and pDC frequencies in healthy controls and COVID-19 patients show. Violin plots depict the percentage of cells in the total population. False detection rate (FDR) of population comparisons for plasmablasts is 4.2×10^−13^ and for pDC 3.1×10^−5^. **E)** UMAP plots of the three immunotypes (BRI n=9, EXI n=6, LAI n=17). **F)** Volcano plots of the pairwise comparison of immunotypes. Positive fold change indicates increase in numerator of ratio, while negative fold change indicates decreases. All patients are from Rotterdam cohort. **G)** Intermediate monocyte 2 frequencies in EXI and LAI patients show. Violin plots depict the percentage of cells in the total population. False detection rate (FDR) of population comparisons depicted in violin plots.

## Discussion

Grouping COVID-19 patients on the basis of severity to understand mechanisms of disease is based on the assumption that one pathophysiological mechanism underlies COVID-19 severity, and that patients respond uniformly to infection and display a similar linear disease progression with time. Using an unbiased analysis that only includes immune parameters can thus provide a more comprehensive mapping of the interplay between the immune response and SARS-CoV-2 virus in patients. Indeed, using this approach, we were able to clearly demarcate immunotypes that predict clinical severity changes and disease outcome. Importantly, the three immunotypes are able to predict disease severity progression, recovery or persistent severe disease. A previous study reported IL-6, IP-10 and IL-10 of first blood draw as predictors of COVID-19 clinical deterioration while antibody responses negatively correlated with deterioration (*7*). However, in that study, 25% of patients that clinically improve still have high levels of IP-10 and would be classified as patients that will worsen (*7*). Similarly, in our study, IL-6 and IL-10 on their own, do not separate well the 3 immunotypes as there is considerable overlap in serum levels. Such overlap is also apparent in antibody responses where BRI and EXI have similar anti-SARS-CoV-2 antibody levels but clearly very different clinical courses. Thus we argue that the combined signature of multiple cytokines and antibodies has more power to predict disease course and outcomes.

The immunotypes described herein cannot be attributed to strictly chronological differences of the duration of infection in patients. Instead of time since infection, the identified immunotypes reflect the variation in individual patient’s kinetics of mounting innate and adaptive immunity to the virus. This notion is supported by the large variation in kinetics with which different hosts mount anti-SARS-CoV-2 immunity (*14*). This variation could be due to host factors such as different host genetics (*15*), epigenetic differences imposed by varying host immunological experiences such as previous pathogen exposures or vaccination (*16*), or potential microbiome differences (*17*). All these factors could lead to varying kinetics of immunity and control of viral loads which can associate with disease severity and clinical course (*18, 19*). Non-host related factors such as dose or route of infection may also play a significant role in immunotype development (*20*). These elements could all influence the rapidity of mounting innate or adaptive immunity or even the duration of different phases of innate immunity. The question arises, whether there are more immunotypes that can be revealed in larger cohorts of patients. Our finding that small subgroups of patients in our cohorts have increased IL-17A or IL-5 in serum, points to additional immunotypes that may have distinct clinical characteristics in acute infection or associate with post-recovery sequelae of long COVID-19. Our study of 138 patients did not have the power to reveal such rarer immunotypes, but larger cohorts may well be able to do so.

The 3 identified immunotypes reflect different pathophysiological mechanisms for COVID-19. Based on serum markers of inflammation such as CRP and d-dimer, BRI and LAI are less inflammatory than EXI. However, LAI exhibits more thrombocytopenia compared to BRI and EXI. Network analysis of the salient association between select pro-inflammatory cytokines, interferons and anti-SARS-CoV-2 antibodies, revealed clear associations of immunotypes with the adaptive immune response, inflammation and the antiviral response (Figure 7). BRI appears to mount an early anti-SARS-CoV-2 antibody response that controls viral replication, and dampens the IFN response and subsequent hyper-inflammation. EXI mounts a vigorous inflammatory response. LAI arises as a result of a delayed adaptive immune response against SARS-CoV-2 that leads to sustained high viral loads accompanied by a type I IFN response. Clearly an impaired type I IFN response is detrimental to the clinical course of COVID-19 as indicated by individuals with defects of IFN signalling and antibodies against IFN (*21-23*). However, increased type I IFN in serum has also been reported in severe COVID-19 patients (*3, 24*) although others have reported low responses (*25*). Such differences may be due to varying patient populations and grouping of different immunotypes. Employing single cell sequencing on just 21 patients, one study reported that anti-SARS-CoV-2 antibodies can hamper type I IFN responses in severe COVID-19 patients (*26*). Although, this could be the case in EXI patients, such results should be interpreted cautiously, as others have reported that antibodies protect from severe COVID-19 (*7*). Additionally, we clearly show that high type I IFN and absence of antibodies in LAI patients associates with disease deterioration and markedly worse outcomes, while BRI patients with high antibodies but no type I IFN do well clinically. The increased IFNα and viral loads in LAI, but their failure to correlate (figure 5D) indicates that in this immunotype their interaction is potentially deregulated. Such correlations can be seen *in vivo* in viral infections such as SIV infection, where uncontrolled viremia positively correlates with type I IFN at the chronic phase, yet negatively correlates at early SIV infection (*27*). On the other hand, type I IFN can induce ACE2 receptor expression (*28*). This can set up a positive feedback loop between viral replication and type I IFN in LAI patients and together with the antiviral effect of type I IFN may set up a complex non-linear interplay between them.

**Figure 7:**
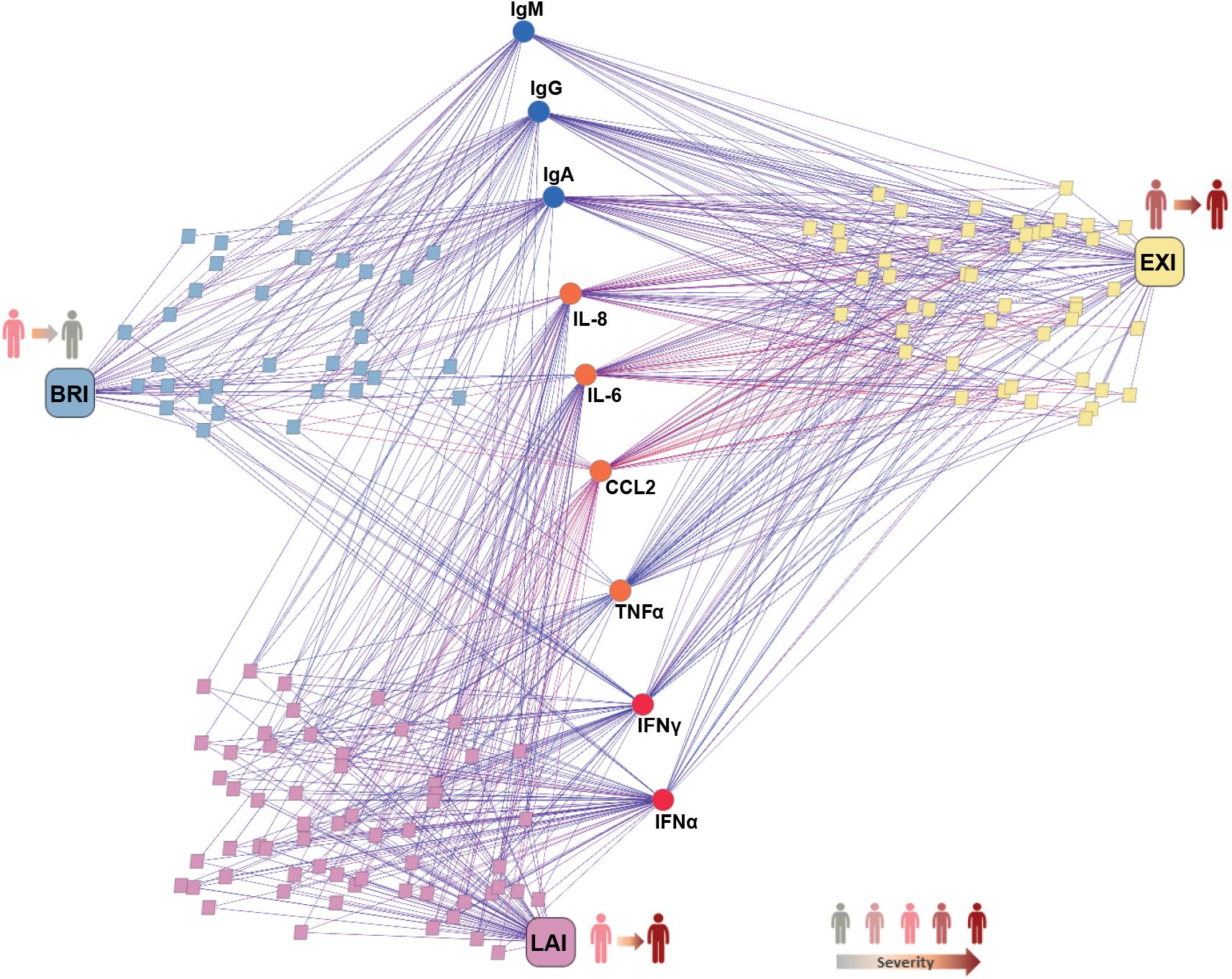
Network analysis of immunotype and molecular marker associations. Schematic representation of the overlapping salient associations across phenotypes with molecular markers shown. Immunotypes BRI, EXI and LAI are shown as rounded rectangles with pastel cyan, yellow, pink respectively. Patient samples are shown in same color, represented by small rectangles and are connected to immunotypes for annotation purposes only. Normalized weighted measurements by the median measurements for each molecular marker seen in a patient sample are represented by colored edges: warmer color signifies higher weight, cooler color signifies lower weight; only the top 1/3 of edges are shown, for clarity. Molecular marker groups are colored by accentuated colors to their respective immunotypes, namely blue, orange, and red. Cross-connections are best seen for CCL2 and to a lesser extent IL-6, connecting mostly EXI with LAI and all three immunotypes, respectively. Antibodies connect to BRI and EXI, while IFN preferentially to LAI. Specificity of immunotypes can be assessed by their average out-degree (d), with BRI being the most specific (d=3.22), followed by LAI (d=3.47) and finally EXI (d=5.37), reflected by the number of edges connecting them to the molecular markers.

We used 40-color spectral flow cytometry to identify the immune cell subset changes associated with COVID-19 infection. We find increased plasmablasts as previously described, but also increases in pro-inflammatory cell types such intermediate monocytes 1 and 2, and pro-inflammatory IgD-CD27-non-conventional memory B cells (*10, 11*), while anti-inflammatory cells such as non-classical monocytes were reduced (*12, 13*). This underscores the tipped balance towards inflammation in patients. Plasmacytoid DC and conventional DC were both reduced in patients and this could impair both anti-viral and adaptive immune responses. Plasmablasts were increased in all 3 immunotypes irrespective if they had antibodies, arguing they are not a good predictor of anti-SARS-CoV-2 immunity. We found only one difference between immunotypes, an increase in EXI patients of pro-inflammatory intermediate monocytes 2 compared to LAI patients. Thus COVID-19 patients had significant cellular variations compared to healthy controls, but such differences between immunotypes were not apparent, indicating that the blood cellular compartment does not directly reflect plasma cytokines that may be produced locally in tissue such as lung.

An exciting prospect of our findings is that classification of COVID-19 patients by immunotype may guide personalized therapeutic strategies. As mentioned, although IL-6 and TNFα were higher in EXI they did not distinguish clearly BRI, EXI and LAI from each other. Treating, therefore, COVID-19 patient based only on IL-6 levels or clinical severity would include all 3 of our immunotypes, and this may explain the relatively small benefit and mixed results of anti-cytokine biologics such as anti-IL-6 antibody (*29-31*). Given the different clinical courses of immunotypes we describe herein, the prediction of which patients will develop a hyperinflammatory syndrome (*32, 33*) may become possible and this would enable pre-emptively blocking key drivers of the pro-inflammatory cytokine network. Our data suggest that LAI patients, which have lower inflammatory cytokines than EXI, for example could benefit most from anti-TNFα and anti-IL-6 antibody therapies as it is well established from sepsis studies that maximal clinical benefit and dampening of cytokine storm is achieved if anti-cytokine treatment is applied before or very early after these cytokines are induced (*34-37*). The potential benefit of prophylactic anti-cytokine treatment for COVID-19 is also suggested from autoimmune patients in which anti-cytokine therapy associates with milder COVID-19 disease (*38-40*). Our, our study also indicates that LAI patients, that have muted antibody responses, would potentially benefit the most from anti-SARS-CoV-2 antibody therapy, as these would mitigate the viral load before a pro-inflammatory cytokine storm settles in. Finally, the observation that very few patients in the BRI immunotype progressed to more severe disease could help hospitals overwhelmed with COVID-19 patients to triage which patients could be transferred to step-down units outside the hospital. Using therefore our 3 immunotypes could guide the formulation of personalized therapies for COVID-19 patients based on mechanistic evidence.

Our study proposes immunotypes that are free from the presumption that clinical classification should dominate the analysis and could be the basis for prospective studies. From a clinical perspective, it is important to identify at an early stage which patients may progress in disease severity as the clinical condition of COVID-19 patients can rapidly deteriorate within days. Here we have identified immunotypes that predict disease progression but also shed light on underlying pathways and suggest biomarkers and therapeutic targets before severe COVID-19 develops. Our current study only addressed the immunotypes in the context of acute COVID-19, but it also points towards employing larger cohort studies that may reveal immunotypes that predict long term post-COVID-19 complications. As immunotype identification requires only serum analysis, these immunotypes can be determined rapidly after patient admission and quickly instruct personalized therapy.

## Material and Methods

### Patients

Rotterdam cohort samples were collected from patients (n=50) participating in the ConCOVID nationwide multicenter open-label randomized clinical trial in the Netherlands. Patients were at least 18 years, admitted to the hospital for COVID-19 and SARS-CoV-2 genome positive by RT-PCR test in the previous 96 hours. Patients entered the study and were sampled at a median of 2 days after hospitalization (IQR: 1, 3,75 days). Healthy controls were age and sex matched. The study was reviewed and approved by the institutional review board of the Erasmus University Medical Center. Written informed consent was obtained from every patient or legal representative. All samples were processed and frozen within 3 hours of bleeding.

Barcelona cohort samples and data from patients included in this study were provided by the HUVH Biobank (PT17/0015/0047), integrated in the Spanish National Biobanks Network and they were processed following standard operating procedures with the appropriate approval of the Ethics and Scientific Committees. All patients entered the study and were sampled on day of hospitalization. Clinical severity was classified according to the WHO 8 point COVID-19 disease severity score (at study inclusion for patients and during hospitalization) in which 0 is no clinical or virological evidence of infection, 1 is no limitation of activities, 2 is limitation of activities, 3 is hospitalized, no oxygen, 4 is oxygen by mask or nasal prongs, 5 is non-invasive ventilation or high-flow oxygen, 6 is intubation and mechanical ventilation, 7 is ventilation and additional organ support (vasopressors, renal replacement therapy, ECMO) and 8 is death (*9*). Clinical characteristics, laboratory measurements and treatments for both cohorts are shown in Supplementary Table 1.

### Cytokine measurements

The cytokines of interest (IL-6, TNFα, IL-1β, IL-8, CCL2, IL-18, IL-10, IL-12, IFNγ, IL-5, IL-17A, IL-2, IL-4, IFNα and TGFβ1) were analyzed using the ELLA Simple Plex system (Protein simple, San Jose, CA). After thawing serum samples on ice, they were centrifuged at 1300xG for 5 minutes at room temperature. Two-fold dilutions were prepared in low protein binding plates according to the manufacturer’s instructions. For TGFβ1, samples were first activated with 1N HCl and then neutralized with 1.2N NaOH/0.5 M HEPES. Subsequently these samples were diluted in a factor 1:15. Diluted samples were loaded into ELLA Simple Plex cartridges and analyzed with the ELLA Simple Plex system.

### Anti-SARS-CoV-2 antibody measurements

Anti-SARS-CoV-2 IgM, IgG and IgA antibodies against nucleocapsid protein (N-protein) were measured in serum by ELISA using COVID-19 IgG ELISA (Tecan, 30177447), COVID-19 IgA ELISA (Tecan, 30177446) and COVID-19 IgM ELISA (Tecan, 30177448) according to the manufacturer’s instructions. Positive cut-off for these ELISAs was 11 units.

### High dimensional flow cytometry

PBMC from patients and controls of the Rotterdam cohort were stained with a 40-color antibody panel and analysed using a 5 laser Aurora spectral flow cytometer (Cytek Biosciences, CA). All samples were stained as described previously (*41*) with the adaptation of including annexin V to exclude dead cells and all buffers contained all buffers contained 2.5mM CaCl2. The cleaning, unsupervised, and statistical inference portions of the flow cytometry analysis were performed using OMIQ (www.omiq.ai). We used the unsupervised analysis methods we previously employed (*41*). The workflow included running flowCut to check for changes in channels over acquisition time, UMAP for dimensionality reduction, flowSOM for clustering, and edgeR for statistical inference. For the statistical comparisons of abundance, the FCS files were subsampled to ensure the same number of events were included per group (either immunotype or disease state).

### Unsupervised hierarchical clustering & Principal Component Analysis

Patient’s clinical and serum data from the Erasmus MC and Barcelona cohort were loaded into R (v.4.0.4) (*42*). Only cytokines positive >20% of patients in both cohorts were used in the analysis. Standardized scores (z-scores) for each cohort were calculated based on the log10 transformed cytokine and antibody levels measured in serum. Unsupervised hierarchical clustering was performed using Ward’s Hierarchical Agglomerative Clustering Method (ward.d2) on the Euclidean distances of the z-scores (*43*). The optimal number of clusters for both cohorts was assigned with the NbClust (v1.0.12) package in R (*44*). NbClust provided 30 indices that determined the best number of clusters for both datasets and proposed the appropriate number of clusters using the majority rule. The majority rule indicated three clusters as optimal for both cohorts. Subsequently, heatmaps were plotted using the R package pheatmap (v1.0.12). Principal Component Analysis (PCA) was performed and visualized based on the z-scores using the R function prcomp.

### Network analysis

Network analysis was performed between immunotypes and select pro-inflammatory cytokines (IL-6, TNFα, IL-8, CCL2) interferons (IFNγ and IFNα) and anti-SARS-CoV-2 IgM, IgG and IgA antibodies. Expression values x per molecular marker i were normalized as x’i=(sign(xi)*ln(abs(xi)+1)/ln2)+min(x) (where x is the difference observed-median), to use as weights for edges in graph layout representations. The first part of this simple equation is a transformation in logarithmic scale, and the latter term is a scaling factor to shift values above zero. For graph analysis and visualization, Biolayout (*45*) was used (see also http://biolayout.org/). Of the total 1380 values/measurements, only the remaining top half (689 edges in total) were used for network analysis to achieve clarity.

### Statistical analysis

Statistical analysis was performed using R (v.4.0.4). Normality of the patients’ data was tested using a Shapiro-Wilk normality test. Statistically significant differences between immunotypes were calculated using multiple Students t-tests for patient’s age and Wilcoxon rank-sum tests for all the other variables. Subsequently, p-values were adjusted using Bonferroni correction. Adjusted p-values lower than 0.05 were considered statistically significant. The number of asterisks indicate the level of significance of p-adjusted values: *p < 0.05, **p < 0.01, ***p < 0.001, and ****p < 0.0001. Non-significant results are not shown in figures. Correlations coefficients between the variables in each cohort were calculated using a Spearman’s rank correlation coefficient and presented in correlograms and DFSO correlations. A correlation was considered statistically significant if the p-value was lower than 0.05. Non-parametric local polynomial regression lines were plotted using the locally estimated scatterplot smoothing (LOESS) method.

## Data availability

The data used in this study are available on request from the corresponding author PDK. The data are not publicly available due to participant privacy/consent.

## Code availability

The R-code used to cluster and the statistical analysis the cytokines, antibodies and clinical data can be freely downloaded from https://bitbucket.org/immunology-emc/covid_severity/src/master/

## Acknowledgments

We would like to thank all the patients who participated in the study. We thank all physicians, medical students, study nurses and trial coordinators in the participating centres. We want to particularly acknowledge the patients and the HUVH Biobank (PT17/0015/0047) integrated in the Spanish National Biobanks Network for their collaboration. Thanks to BMW, the Netherlands for providing two free-of-charge vehicles for sample collection. This work was supported by Health Holland LSHM20056 grant (PDK), and in part supported by the Erasmus foundation (BJAR), grant PI20/00416 from the Instituto de Salud Carlos III (RPB) and the European Regional Development Fund (ERDF) (RPB).

## Author contributions

Y.M.M., T.J.S., R.R., M.W.J.S., D.A.S., C.H.K., M.D.C.E., M.v.M., I.B.H., M.Z., L.L., H.d.W., C.A.O., M.E.P.W., T.A., D.A.L., T.v.W., M.C.J., G.K., S.v.B., C.R., B.J.A.R., M.H.G., R.P.B., contributed to data acquisition and analysis. D.G.B.G. and H.J.G.vd.W. performed bioinformatics and statistical analysis. P.D.K. conceived and designed the work, and wrote the manuscript. All authors reviewed and edited the manuscript.

## Materials & Correspondence

Address all correspondence and material requests to P.D.K. at

---

**Supplementary figure S1:**
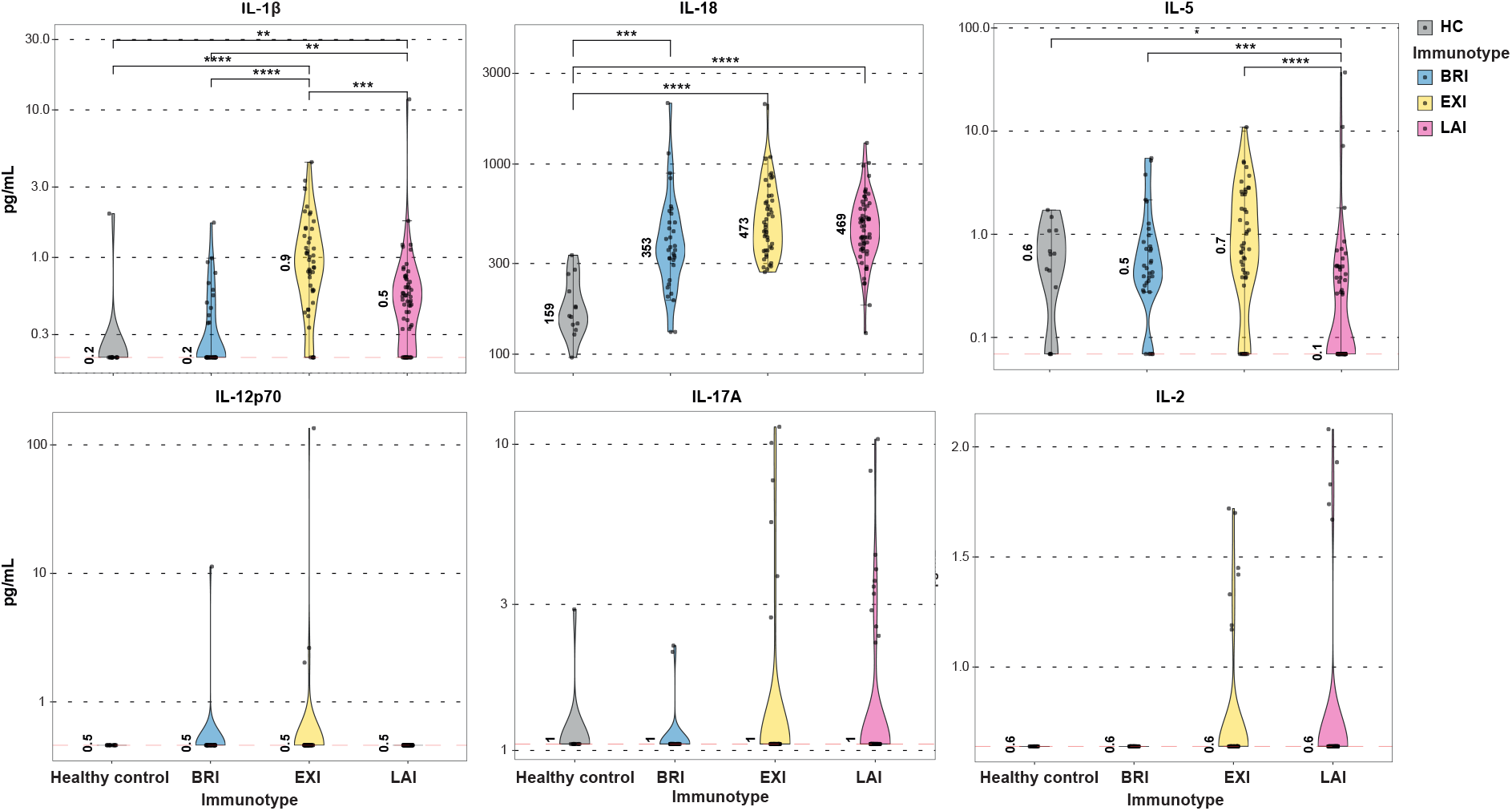
Violin plots depicting serum levels for IL-1β, IL-18, IL-5, IL-12, IL-17A and IL-2 in the patients of different immunotypes and healthy controls. Median values are in numbers and Wilcoxon rank-sum tests with Bonferroni correction were applied for each measurement. Red dashed line indicates assay Limit of Detection. *p < 0.05, **p < 0.01, ***p < 0.001, and ****p < 0.0001.

**Supplementary figure S2:**
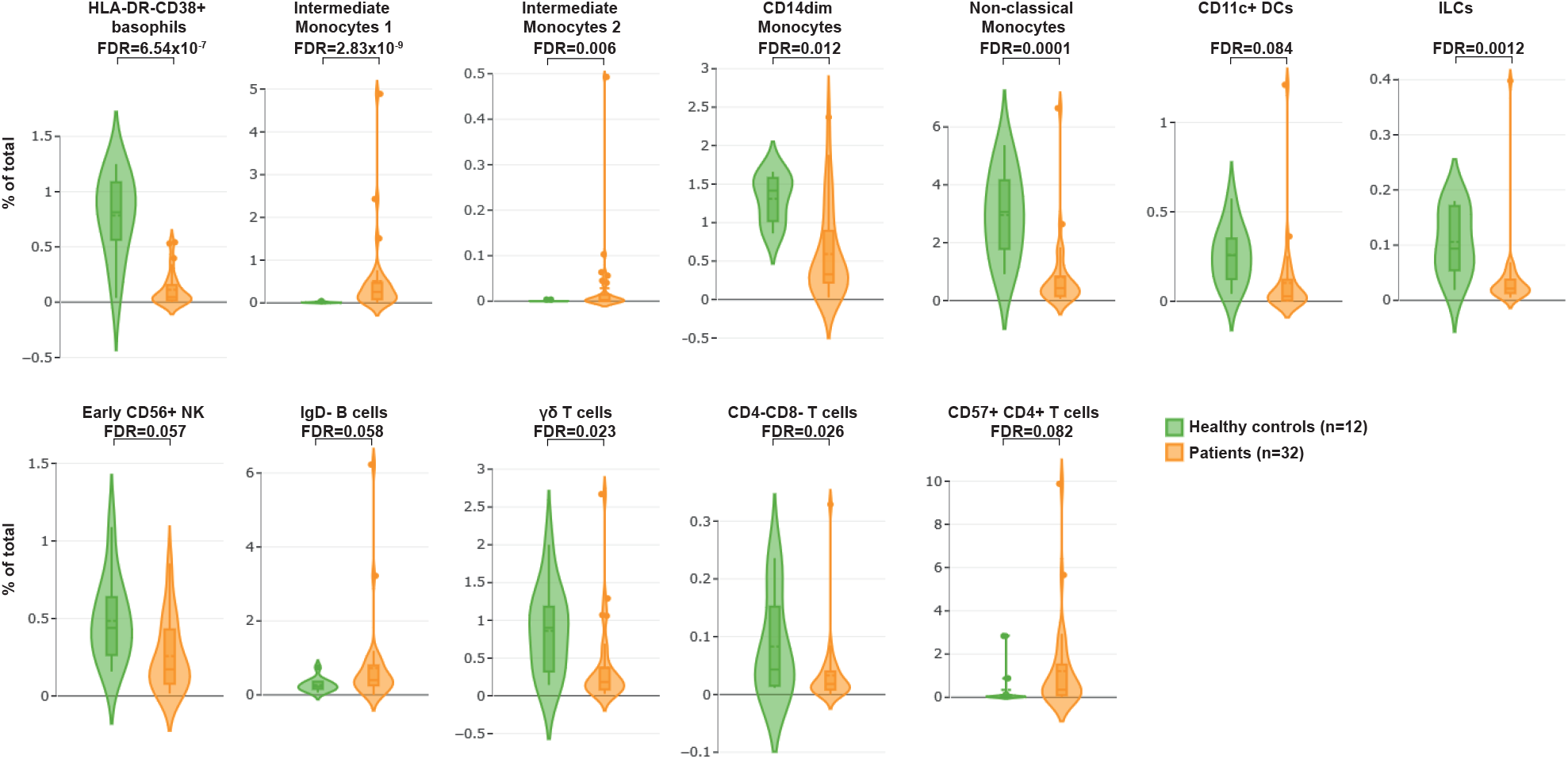
COVID-19 patient (n=35) and healthy control (n=12) peripheral blood cellular populations that were identified by high dimensional 40-color flow cytometry to be significantly different are shown (FDR<0.1). Violin plots show the range of differentially expressed blood populations and depict the percentage of cells in the total population.

**Supplementary Table 1.** Clinical and laboratory characteristics of Rotterdam discovery and Barcelona validation cohorts. The clinical characteristics and laboratory measurements of Rotterdam discovery cohort, Barcelona validation cohort and the combination of both shown.

|  | Level | Overall | Barcelona | Rotterdam |
| --- | --- | --- | --- | --- |
| **N** |  | 138 | 88 | 50 |
| **Immunotype** | BRI | 33 (23.9%) | 19 (21.6%) | 14 (28.0%) |
|  | EXI | 46 (33.3%) | 33 (37.5%) | 13 (26.0%) |
|  | LAI | 59 (42.8%) | 36 (40.9%) | 23 (46.0%) |
| **Gender** | male | 91 (65.9%) | 58 (65.9%) | 33 (66.0%) |
| **Age** | years | 62 (54 – 70) | 61 (50 – 70) | 63 (57.25 – 69) |
| **Days from symptom onset** |  | 8 (6 – 12)<br>[n=134] | 8 (6 – 10)<br>[n=84] | 9 (6 – 14.75) |
| **WHO8-point score at study entry** | 3 | 38 (27.5%) | 34 (38.6%) | 4 (8.0%) |
|  | 4 | 60 (43.5%) | 31 (35.2%) | 29 (58.0%) |
|  | 5 | 20 (14.5%) | 12 (13.6%) | 8 (16.0%) |
|  | 6 | 12 (8.7%) | 10 (11.4%) | 2 (4.0%) |
|  | 7 | 8 (5.8%) | 1 (1.1%) | 7 (14.0%) |
| **Obesity** | yes | 43 (31.2%) | 20 (22.7%) | 23 (46.0%) |
|  | NA | 6 (4.3%) | 2 (2.3%) | 4 (8.0%) |
| **Diabetes mellitus** | yes | 32 (23.2%) | 18 (20.5%) | 14 (28.0%) |
|  | NA | 2 (1.4%) | 2 (2.3%) | 0 (0.0%) |
| **Heart disease** | yes | 48 (34.8%) | 35 (39.8%) | 13 (26.0%) |
|  | NA | 2 (1.4%) | 2 (2.3%) | 0 (0.0%) |
| **Lung disease** | yes | 30 (21.7%) | 15 (17.0%) | 15 (30.0%) |
|  | NA | 2 (1.4%) | 2 (2.3%) | 0 (0.0%) |
| **Kidney disease** | yes | 5 (3.6%) | 3 (3.4%) | 2 (4.0%) |
|  | NA | 2 (1.4%) | 2 (2.3%) | 0 (0.0%) |
| **Liver disease** | yes | 6 (4.3%) | 5 (5.7%) | 1 (2.0%) |
|  | NA | 2 (1.4%) | 2 (2.3%) | 0 (0.0%) |
| **Cancer** | yes | 7 (5.1%) | 3 (3.4%) | 4 (8.0%) |
|  | NA | 2 (1.4%) | 2 (2.3%) | 0 (0.0%) |
| **Fever (>38 °C)** | yes | 91 (65.9%) | 68 (77.3%) | 23 (46.0%) |
|  | NA | 2 (1.4%) | 2 (2.3%) | 0 (0.0%) |
| **C-reactive protein (mg/L)** |  | 102 (68 - 181)<br>[n=127] | 116 (73 - 192)<br>[n=78] | 96 (52 - 151)<br>[n=49] |
| **Ferritin (µg/L)** |  | 666 (382 - 1101)<br>[n=116] | 598 (314 - 1034)<br>[n=72] | 738 (487 - 1171)<br>[n=44] |
| **Lactate dehydrogenase (U/L)** |  | 369 (293 - 472)<br>[n=121] | 369 (296 - 474)<br>[n=72] | 373 (286 - 461)<br>[n=49] |
| **Alanine aminotransferase (U/L)** |  | 29 (19 - 54)<br>[n=137] | 32 (18 - 54)<br>[n=88] | 25 (21 - 54)<br>[n=49] |
| **Bilirubin (µmol/L)** |  | 9.8 (7.1 - 13.0)<br>[n=122] | 10.1 (7.8 - 13.2)<br>[n=76] | 8.5 (6.0 - 12.0)<br>[n=46] |
| **D-dimer (mg/L)** |  | 0.40 (0.24 - 0.98)<br>[n=120] | 0.30 (0.19 - 0.48)<br>[n=79] | 0.97 (0.60 - 1.49)<br>[n=41] |
| **Hemoglobin (mmol/L)** |  | 8.30 (7.51 - 9.06)<br>[n=136] | 8.66 (7.88 - 9.18)<br>[n=86] | 7.65 (6.60 - 8.30)<br>[n=50] |
| **Leukocytes (x10<sup>9</sup>/L)** |  | 7.45 (5.90 - 9.84)<br>[n=136] | 7.62 (6.11 - 9.87)<br>[n=86] | 7.00 (5.20 - 9.78)<br>[n=50] |
| **Neutrophils (x10<sup>9</sup>/L)** |  | 5.70 (4.16 - 8.01)<br>[n=133] | 6.02 (4.60 - 8.03)<br>[n=86] | 5.10 (3.70 - 7.70)<br>[n=47] |
| **Lymphocytes (x10<sup>9</sup>/L)** |  | 1.02 (0.83 - 1.41)<br>[n=133] | 1.02 (0.83 - 1.38)<br>[n=86] | 1.00 (0.80 - 1.50)<br>[n=47] |
| **Monocytes (x10<sup>9</sup>/L)** |  | 0.45 (0.30 - 0.64)<br>[n=133] | 0.48 (0.33 - 0.66)<br>[n=86] | 0.40 (0.29 - 0.60)<br>[n=47] |
| **Thrombocytes (x10<sup>9</sup>/L)** |  | 216 (163 - 294) | 209 (166 - 289) | 229 (151 - 298) |

|  |  | [n=135] | [n=86] | [n=49] |
| --- | --- | --- | --- | --- |
| **Tocilizumab before study entry** | yes | 7 (5.1%) | 0 (0.0%) | 7 (14.0%) |
|  | NA | 0 (0.0%) | 0 (0.0%) | 0 (0.0%) |
| **Tocilizumab after study entry** | yes | 2 (1.4%) | 0 (0.0%) | 2 (4.0%) |
|  | NA | 88 (63.8%) | 88 (100.0%) | 0 (0.0%) |
| **Corticosteroids after study entry** | yes | 10 (7.2%) | 0 (0.0%) | 10 (20.0%) |
|  | NA | 88 (63.8%) | 88 (100.0%) | 0 (0.0%) |
| **Convalescent plasma after study entry** | yes | 29 (21.0%) | 0 (0.0%) | 29 (58.0%) |
|  | NA | 88 (63.8%) | 88 (100.0%) | 0 (0.0%) |
| Data are n (%) or median (IQR). Complete data was available for the continuous values shown if not stated otherwise [n=] ; NA not available. |  |  |  |  |

**Supplementary Table 2.** Clinical and laboratory characteristics of BRI, EXI and LAI immunotypes. The clinical characteristics and laboratory measurements of patients in each immunotype group are presented.

| Supplementary Table 2. Clinical and laboratory characteristics of immunotypes BRI, EXI and LAI. |  |  |  |  |  |
| --- | --- | --- | --- | --- | --- |
|  | Level | BRI | EXI | LAI | P-value |
| N |  | 33 | 46 | 59 |  |
| Cohort | Barcelona | 19 (57.6%) | 33 (71.7%) | 36 (61.0%) | 0.367 |
|  | Rotterdam | 14 (42.4%) | 13 (28.3%) | 23 (39.0%) |  |
| Gender | male | 15 (45.5%) | 36 (78.3%) | 40 (67.8%) | 0.009 |
| Age | years | 57 (49 – 64) | 63 (57 – 69.75) | 65 (52.50 – 71) | 0.059 |
| Days from symptom onset |  | 10 (8 – 13) | 10 (6.50 – 14)<br>[n=43] | 7 (5 – 10)<br>[n=58] | 0.002 |
| WHO8-point score at study entry | 3 | 17 (51.5%) | 5 (10.9%) | 16 (27.1%) | 0.002 |
|  | 4 | 11 (33.3%) | 20 (43.5%) | 29 (49.2%) |  |
|  | 5 | 3 (9.1%) | 8 (17.4%) | 9 (15.3%) |  |
|  | 6 | 0 (0.0%) | 8 (17.4%) | 4 (6.8%) |  |
|  | 7 | 2 (6.1%) | 5 (10.9%) | 1 (1.7%) |  |
| Obesity | yes | 6 (18.2%) | 13 (28.3%) | 24 (40.7%) | 0.192 |
|  | NA | 1 (3.0%) | 3 (6.5%) | 2 (3.4%) |  |
| Diabetes mellitus | yes | 4 (12.1%) | 11 (23.9%) | 17 (28.8%) | 0.113 |
|  | NA | 0 (0.0%) | 2 (4.3%) | 0 (0.0%) |  |
| Heart disease | yes | 6 (18.2%) | 20 (43.5%) | 22 (37.3%) | 0.034 |
|  | NA | 0 (0.0%) | 2 (4.3%) | 0 (0.0%) |  |
| Lung disease | yes | 8 (24.2%) | 11 (23.9%) | 11 (18.6%) | 0.311 |
|  | NA | 0 (0.0%) | 2 (4.3%) | 0 (0.0%) |  |
| Kidney disease | yes | 0 (0.0%) | 4 (8.7%) | 1 (1.7%) | 0.049 |
|  | NA | 0 (0.0%) | 2 (4.3%) | 0 (0.0%) |  |
| Liver disease | yes | 1 (3.0%) | 3 (6.5%) | 2 (3.4%) | 0.293 |
|  | NA | 0 (0.0%) | 2 (4.3%) | 0 (0.0%) |  |
| Cancer | yes | 2 (6.1%) | 1 (2.2%) | 4 (6.8%) | 0.271 |
|  | NA | 0 (0.0%) | 2 (4.3%) | 0 (0.0%) |  |
| Fever (>38 °C) | yes | 15 (45.5%) | 29 (63.0%) | 47 (79.7%) | 0.004 |
|  | NA | 0 (0.0%) | 2 (4.3%) | 0 (0.0%) |  |
| C-reactive protein (mg/L) |  | 82 (39 – 125)<br>[n=32] | 159 (86 – 245)<br>[n=44] | 94 (59 – 152)<br>[n=51] | <0.001 |
| Ferritin (µg/L) |  | 434 (278 – 676)<br>[n=32] | 887 (648 – 1883)<br>[n=38] | 605 (313 – 1052)<br>[n=46] | <0.001 |
| Lactate dehydrogenase (U/L) |  | 320 (257 – 381)<br>[n=32] | 453 (362 – 599)<br>[n=39] | 341 (296 – 454)<br>[n=50] | <0.001 |
| Alanine aminotransferase (U/L) |  | 25 (18 – 54) | 31 (18 – 57)<br>[n=46] | 29 (21 – 53)<br>[n=58] | 0.725 |
| Bilirubin (µmol/L) |  | 8.9 (6.9 – 13.0)<br>[n=31] | 11.4 (8.0 – 14.8)<br>[n=44] | 9.0 (6.0 – 12.2)<br>[n=47] | 0.1 |
| D-dimer (mg/L) |  | 0.39 (0.30 – 0.61)<br>[n=29] | 0.89 (0.32 – 2.22)<br>[n=39] | 0.30 (0.18 – 0.62)<br>[n=52] | <0.001 |
| Hemoglobin (mmol/L) |  | 8.01 (7.45 – 8.69) | 8.30 (7.60 – 9.05)<br>[n=44] | 8.38 (7.70 – 9.12) | 0.352 |
| Leukocytes (x10<sup>9</sup>/L) |  | 7.35 (6.10 – 9.30) | 9.70 (7.10 – 12.33)<br>[n=44] | 6.50 (4.84 – 8.06) | <0.001 |
| Neutrophils (x10<sup>9</sup>/L) |  | 5.11 (4.16 – 7.69) | 7.78 (5.69 – 10.05)<br>[n=44] | 4.78 (3.61 – 6.47)<br>[n=56] | <0.001 |
| Lymphocytes (x10<sup>9</sup>/L) |  | 1.40 (0.90 – 1.50) | 0.99 (0.82 – 1.26)<br>[n=44] | 1.00 (0.80 – 1.28)<br>[n=56] | 0.067 |
| Monocytes (x10<sup>9</sup>/L) |  | 0.48 (0.40 – 0.68) | 0.50 (0.30 – 0.72)<br>[n=44] | 0.38 (0.27 – 0.58)<br>[n=56] | 0.008 |
| Thrombocytes (x10<sup>9</sup>/L) |  | 280 (208 – 343) | 274 (176 – 320)<br>[n=44] | 182 (152 – 229)<br>[n=58] | <0.001 |
| **Tocilizumab before study entry** | yes | 0 (0.0%) | 6 (13.0%) | 1 (1.7%) | 0.01 |
|  | NA | 0 (0.0%) | 0 (0.0%) | 0 (0.0%) |  |
| **Tocilizumab after study entry** | yes | 1 (3.0%) | 0 (0.0%) | 1 (1.7%) | 0.582 |
|  | NA | 19 (57.6%) | 33 (71.7%) | 36 (61.0%) |  |
| **Corticosteroids after study entry** | yes | 3 (9.1%) | 4 (8.7%) | 3 (5.1%) | 0.476 |
|  | NA | 19 (57.6%) | 33 (71.7%) | 36 (61.0%) |  |
| **Convalescent plasma after study entry** | yes | 9 (27.3%) | 5 (10.9%) | 15 (25.4%) | 0.356 |
|  | NA | 19 (57.6%) | 33 (71.7%) | 36 (61.0%) |  |
| Data are n (%) or median (IQR). Complete data was available for the continuous values shown if not stated otherwise [n=]; NA not available. Significance between immunotypes for categorical variables determined by Fisher's exact test and for continuous variables by Kruskal-Wallis Rank Sum Test. |  |  |  |  |  |

**Supplementary Table 3.**
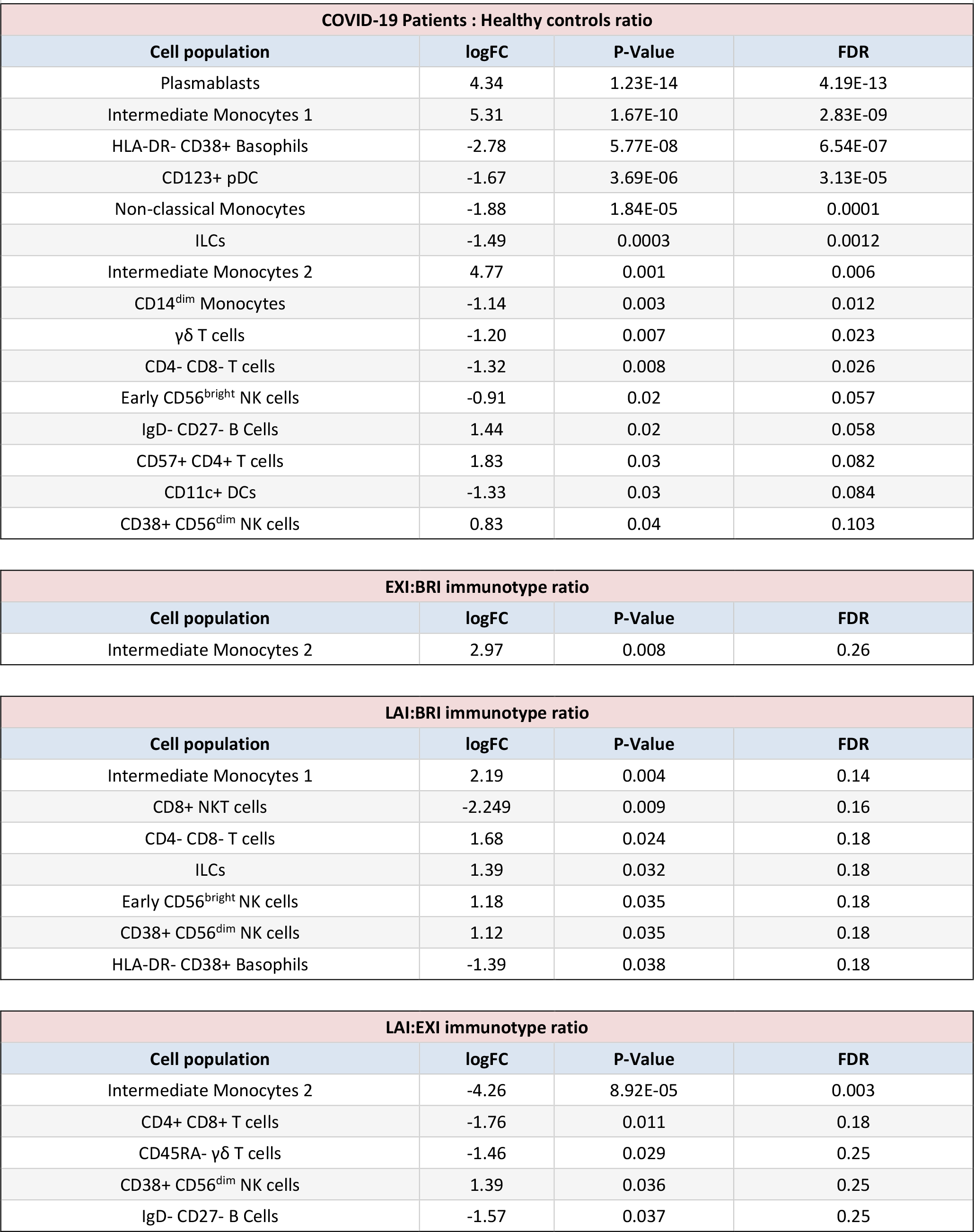
High dimensional flow cytometry identified differentially expressed cellular populations. The peripheral blood cellular populations identified by high dimensional 40-color flow cytometry from indicated pairwise comparisons are shown. Comparisons between COVID-19 patients (n=32) and healthy controls (n=12), and pairwise comparisons of immunotypes (BRI n=9, EXI n=6 and LAI n=17) shown. Positive logFC indicates numerator of ratio is increased while negative values indicate a decrease. LogFC indicate log2 fold change. Comparisons with FDR<0.1 are considered to be significantly different. All population comparisons with a p<0.05 are shown.

| COVID-19 Patients : Healthy controls ratio |  |  |  |
| --- | --- | --- | --- |
| Cell population | logFC | P-Value | FDR |
| Plasmablasts | 4.34 | 1.23E-14 | 4.19E-13 |
| Intermediate Monocytes 1 | 5.31 | 1.67E-10 | 2.83E-09 |
| HLA-DR- CD38+ Basophils | -2.78 | 5.77E-08 | 6.54E-07 |
| CD123+ pDC | -1.67 | 3.69E-06 | 3.13E-05 |
| Non-classical Monocytes | -1.88 | 1.84E-05 | 0.0001 |
| ILCs | -1.49 | 0.0003 | 0.0012 |
| Intermediate Monocytes 2 | 4.77 | 0.001 | 0.006 |
| CD14<sup>dim</sup> Monocytes | -1.14 | 0.003 | 0.012 |
| γδ T cells | -1.20 | 0.007 | 0.023 |
| CD4- CD8- T cells | -1.32 | 0.008 | 0.026 |
| Early CD56<sup>bright</sup> NK cells | -0.91 | 0.02 | 0.057 |
| IgD- CD27- B Cells | 1.44 | 0.02 | 0.058 |
| CD57+ CD4+ T cells | 1.83 | 0.03 | 0.082 |
| CD11c+ DCs | -1.33 | 0.03 | 0.084 |
| CD38+ CD56<sup>dim</sup> NK cells | 0.83 | 0.04 | 0.103 |

| EXI:BRI immunotype ratio |  |  |  |
| --- | --- | --- | --- |
| Cell population | logFC | P-Value | FDR |
| Intermediate Monocytes 2 | 2.97 | 0.008 | 0.26 |

| LAI:BRI immunotype ratio |  |  |  |
| --- | --- | --- | --- |
| Cell population | logFC | P-Value | FDR |
| Intermediate Monocytes 1 | 2.19 | 0.004 | 0.14 |
| CD8+ NKT cells | -2.249 | 0.009 | 0.16 |
| CD4- CD8- T cells | 1.68 | 0.024 | 0.18 |
| ILCs | 1.39 | 0.032 | 0.18 |
| Early CD56<sup>bright</sup> NK cells | 1.18 | 0.035 | 0.18 |
| CD38+ CD56<sup>dim</sup> NK cells | 1.12 | 0.035 | 0.18 |
| HLA-DR- CD38+ Basophils | -1.39 | 0.038 | 0.18 |

**Supplementary Table 3. High dimensional flow cytometry identified differentially expressed cellular populations.**
| LAI:EXI immunotype ratio |  |  |  |
| --- | --- | --- | --- |
| Cell population | logFC | P-Value | FDR |
| Intermediate Monocytes 2 | -4.26 | 8.92E-05 | 0.003 |
| CD4+ CD8+ T cells | -1.76 | 0.011 | 0.18 |
| CD45RA- γδ T cells | -1.46 | 0.029 | 0.25 |
| CD38+ CD56<sup>dim</sup> NK cells | 1.39 | 0.036 | 0.25 |
| IgD- CD27- B Cells | -1.57 | 0.037 | 0.25 |

